# Sentiment analysis of employees and COVID-19 vaccine hesitancy at workplace

**DOI:** 10.1101/2025.01.30.25321414

**Authors:** Ikpe Justice Akpan, Teai Warner, Mbuotidem Peters

## Abstract

Vaccination is a potent means to combat the spread of infectious disease epidemics or pandemics, such as the COVID-19 pandemic. However, getting sufficient people to accept the vaccine and achieve herd immunity remains a significant challenge. This study evaluates preschool workers’ sentiments and challenges during in-person schooling amidst the COVID-19 pandemic and attitudes toward COVID-19 vaccination through a survey. The study surveyed preschool workers as part of a consulting project. Workers’ sentiments are analyzed using Azure Machine Learning (AML) data analytic method, statistical analysis, and theoretical evaluation. The results show that no association exists between employee job category (teachers versus support staff) and COVID-19 vaccine hesitancy. However, fewer teaching staff were hesitant to take the COVID-19 vaccine than the support staff (46% < 50%), but the difference was not significant [Chi-Square (χ2) = 0.009; p>0.05]. The overall sentiments of all preschool workers showed 46%, indicating low neutrality, implying hesitancy toward the COVID-19 vaccine. Preschool workers who felt optimistic about the vaccine’s potency cited scientific reasons; those with neutral sentiments needed further information and assurances about the side effects, indicating that appropriate health education can sway more people positively towards accepting the vaccine; people with negative sentiments displayed distrust, fear, personal beliefs, and misinformation. The results also highlight the need to educate all workers on vaccine potency despite the levels of education. Regardless of academic qualifications, any uninformed person can fall prey to misinformation and conspiracy theories. Proper health education helps people make informed decisions.

## 1. Introduction

The coronavirus disease that broke out in December 2019 (COVID-19) infected and killed millions of people worldwide and caused a significant devastating impact on workers [1]. The community lockdowns that ensued to limit the pandemic spread (through infection and fatality) caused substantial disruptions in communities and workplaces, impacting social and work life [2-4]. Several studies report that the ravaging impacts of COVID-19 generated fear of safety, work-related stress, and mental health problems in the workplace, especially in the healthcare and education sectors and in the broader society [5-7]. The workers, students, and families raised several pandemic-related concerns in schools. In the USA, where this study took place, the National Head Start Association (NHSA) also identified these struggles, including e-learning challenges, psychological problems, high unemployment, and family instability [3,8,9]. NHSA is an organization that supervises special preschools (known as Early Head Start), which serves students from low-income backgrounds. The identified challenges further caused domestic abuse and neglect during COVID-19 in some families [9-12]. Also, the parents of preschool students expressed concerns about children’s academic and social well-being while attending school [13,14], while teachers seemed unsure about ways to stay safe while teaching the children during the pandemic amidst limited resources [14,15].

As the above challenges mounted, preschools, which undoubtedly operate with smaller budgets than high schools and tertiary institutions, struggled to open during the pandemic for several reasons. First, there was a shortage of personal protective equipment (PPE) due to supply chain problems [16-17]. PPEs (e.g., face covering) were part of the public health guidelines intended to keep people safe based on the CDC guidelines [18-20]. However, other public health safety measures, such as social distancing, were strictly and successfully observed at the preschool [3,17]. Secondly, there were employee shortages due to isolation or quarantine. Based on the public health safety requirements, employees at risk of COVID-19 or those exposed to such risks were isolated or quarantined [18,20-22]. These prevailing circumstances at work frustrated employees, students, and their parents. Also, the preschool operated with fewer students due to the social distancing guidelines recommended by the public health authority [19,21].

Given the above workplace challenges, the race to produce and administer the COVID-19 vaccine intensified as one of the most effective ways to combat the deadly pandemic and return to normalcy [23]. Several studies provide evidence that vaccine is one of the most effective ways to control and limit the spread of the coronavirus disease, reduce mortality, and prevent severe illness in the event of contracting the disease [23,24]. For example, a vaccinated person is less likely to develop a severe health condition if infected by COVID-19 [24,25]. Also, COVID-19 vaccination can help combat the spread of the pandemic, especially when combined with other public health measures and reaching herd immunity [20,25-27].

Considering the potential of accelerated transmission and spread of the deadly coronavirus disease in academic communities, schools, colleges, and universities were closed worldwide, especially during the first and second waves of the pandemic [28-30]. Educational institutions, especially in advanced economies, quickly adopted/improvised technologies and moved learning to the virtual space through e-learning implementation worldwide, which became the new standard [30-33]. In societies where technologies to implement virtual learning were deficient, schools were shut temporarily to limit the spread of SARS-CoV-2, leading to learning loss, especially in less developed countries [34,35]. Meanwhile, efforts to discover and develop a COVID-19 vaccine intensified as researchers, scientists, and corporate organizations scouted effective solutions against the deadly pandemic [35,36]. The scientific efforts towards the COVID-19 vaccine yielded fruits as it was discovered and produced in record time [37,38]. The next challenge was to get many people in the academic communities to take the vaccine to achieve herd immunity, which was the most effective and practical solution to safe reopening and a return to traditional in-person schooling after the initial closures [39].

This study examined the general sentiments of workers in the preschool organization and gathered employees’ opinions regarding addressing potential COVID-19 vaccine hesitancy. The survey of workers in this study formed part of a consulting project that evaluated modalities for safe reopening, formulating strategies for a safe return to traditional face-to-face learning, and managing the challenging situations caused by the pandemic [3,17].

Although several studies identified factors influencing COVID-19 vaccine hesitancy, those research does not involve workers in early learning institutions [40-42]. This study fills that gap by examining preschool employees’ sentiments and analyzing the factors that impact COVID-19 vaccine hesitancy in educational organizations. Specifically, we address the following research objectives:

i. Analyze preschool workers’ general sentiments about COVID-19 vaccination intentions (RO1).
ii. Evaluate the association between employee job category (teachers versus support/office staff) and COVID-19 vaccine hesitancy (RO2).
iii. Examine the factors associated with the COVID-19 vaccination hesitancy among the preschool workers (RO3).

## 2. Materials and Method

### 2.1 The Sample

This study surveyed all the employees at a preschool, including teaching and support at the institution, which operates as a private non-profit organization offering education and daycare. The teaching staff included fully qualified teachers and those in the support-teachers role. The non-teaching employees occupied different roles, including school bus drivers, office administration, family support or advocates, and other positions. Some workers performed multiple roles, such as bus driver and office support staff.

### 2.2 Questionnaire design and administration

This survey was conducted in collaboration with a preschool organization in a mid-west USA county, which approved the study and administered the questionnaire. The survey was part of a consulting project commissioned to help identify survival strategies and safe reopening of the early learning educational institution after the initial closure due to the COVID-19 pandemic. The research consultants created the anonymous questionnaires in collaboration with the preschool directors, who edited and approved the survey materials based on their internal policies. The preschool also administered the survey to the employees as respondents. Participation in the survey was entirely optional. Based on the preschool’s privacy policy, the authors did not have direct access to the respondents’ contact information. Therefore, the preschool admin staff administered the questionnaire using ‘survey-monkey.com,’ a popular online survey platform quite familiar to many researchers [43]. Ensuring anonymity motivated the respondents to offer honest and accurate responses to the questions [44]. As a way of ensuring the anonymity of respondents, the questionnaire did not contain any attributes capable of identifying the participants, e.g., the survey did not collect information about the sex of the respondents. The questionnaire was largely multiple-choice and open-ended questions. The survey occurred between February and March 2021. There were thirty (30) usable responses.

### 2.3 Factors Influencing Employees’ Vaccine Hesitancy

Recent studies identified employees’ level of education and type of employment (e.g., essential workers or frontline employees) can impact COVID-19 vaccination hesitancy [45]. The respondents in our study have diverse backgrounds in terms of academic qualifications, ranging from ‘no college’ to workers with college degrees. The teachers are also required to have appropriate teaching certification. Similarly, some support staff positions, such as the "family advocates," also require a college degree. However, some staff, such as bus drivers, do not need a college degree, although they work in close physical contact with the students.

Respondents were asked about plans and willingness to take vaccination if a COVID-19 vaccine if/when offered and presented with three options to choose from, including "yes," "no," and "unsure." Another critical question asked the survey participants to enumerate or explain the reasons for their decisions (accepting vaccination or being hesitant). The survey results were coded as factors for or against taking the COVID-19 vaccine when offered. Previous studies adopted a similar strategy to evaluate past vaccine hesitancy [45].

### 2.4 Methods for Analyzing Results

The data collected from the survey are mainly categorical and textual. The nominal data from the categorical variables are analyzed using non-parametric statistics known as the Chi-Square method [46]. The process involves identifying and categorizing outputs and themes using cross-tabulations and comparing frequencies. Other analyses involve comparing the outcomes based on the respondents’ work roles. The classification includes teaching staff (teachers and classroom assistants) and non-teaching support workers (family advocates, bus drivers, office admin, and more). For example, when analyzing employees’ perceptions about taking the COVID-19 vaccines, the results compare the respondents’ positions based on work roles unless the number of responses for the categories is considered too small for any meaningful analysis.

The second aspect of the study involves collecting and analyzing textual data evaluated using sentiment analysis based on the Azure machine learning (AML) technique. AML is a cloud-based application offered by Microsoft (MS) that enables analysts and data model developers to build, train, and deploy ML models covering the entire ML lifecycle (including data preparation, model training, and monitoring) [47,48]. The AML application can be embedded in R-studio, Python, or MS Excel [47,48]. In this study, AML was deployed in the MS Excel environment, one of the software components of the MS Office 365 application, and used to analyze the perceptions and sentiments of the survey respondents, otherwise known as sentiment analysis or opinions mining [46]. It is a natural language processing technique that evaluates the respondent’s perception or position about a topic of interest based on subjective reasoning [49]. Opinions or perceptions are often subjective and can be categorized into one of three options, namely, positive, neutral, or negative perceptions [17,48,49].

## 3. Results and Discussion

### 3.1 Respondents’ Work Roles

As indicated earlier, the survey questionnaire was anonymous. It generated thirty (30) usable responses. The workers’ job roles (Figure 1) were as follows: teachers (17%), support teachers (17%), Office-Admin Staff (22%), bus drivers (6%) and family advocates (19%). Some of the employees identified more than one job category. For example, one employee served as Special Need Assistant and Office-Admin Staff, and another as a Bus Driver and classroom assistant. The largest single category was "Office-Admin Staff," followed by the "family advocates" (22% and 19%) of respondents.

**Figure 1.**
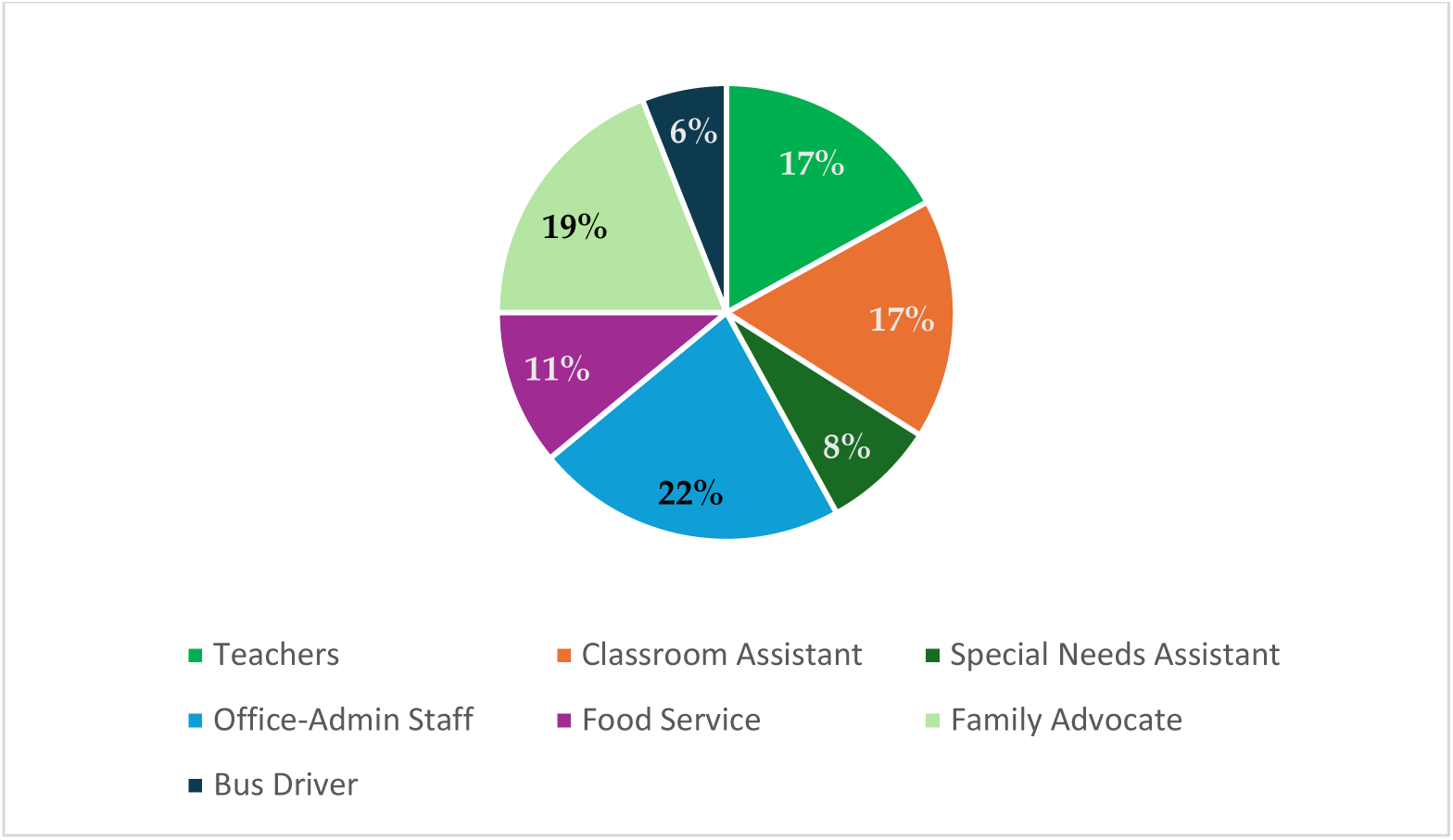
Employee respondents based on job roles.

### 3.2 Workers’ Sentiments About COVID-19 Vaccination

This Section presents the results of workers’ sentiments towards COVID-19 vaccination analyzed using the AML described earlier [48,49]. These results help to address the first research objective (RO1) listed in the earlier Section. The opinions or sentiments were obtained as textual data originated from open-ended questions that constitute respondents’ perceptions and sentiments towards the COVID-19 vaccination as a public health safety measure. The survey asked the respondents about their general opinions about the COVID-19 vaccine and their intention to take the COVID-19 vaccination if/when offered. The responses for this question were textual data, which were coded into eighteen (18) positive, negative, and neutral sentiments, perceptions, and opinions (Table 1). The result shows that seven (7) of the sentiments were portrayed by both teaching staff and non-teaching employees (positive: 4; negative: 3; neutral: 0).

**Table 1.** Employees’ sentiments/reasons for and against COVID-19 vaccination decision.

| Workers' Sentiments/Perceptions | Non-Teaching Staff | Teaching Staff | Sentiment Analysis Outcomes Using Azure Machine Learning |  |  |
| --- | --- | --- | --- | --- | --- |
|  |  |  | Outcome | Score | Percent |
| The COVID-19 vaccine is too new, I cannot trust it. | X | X | Negative | 0.345 | 35% |
| I will take the vaccine so that I can visit family and friends again. | X | X | Positive | 0.697 | 70% |
| I will take vaccine to protect myself, family, and others. | X | X | Positive | 0.692 | 69% |
| I will accept the COVID-19 vaccine so that we can reach herd immunity and stop the pandemic. | X | X | Positive | 0.731 | 73% |
| I will take the vaccine to stay safe and healthy. | X | - | Positive | 0.661 | 66% |
| I cannot take the COVID-19 now because I am trying to get pregnant. | X | X | Negative | 0.134 | 13% |
| I want to observe the vaccine outcomes in other people before I can consider taking it. | - | X | Negative | 0.357 | 36% |
| I am nervous about the COVID-19 vaccine side effects. | X | - | Neutral | 0.522 | 52% |
| I need more information about the COVID-19 vaccine before I decide. | X | X | Negative | 0.349 | 35% |
| I will take the vaccine to help resume normal life. | X | X | Positive | 0.641 | 64% |
| I will take the COVID vaccine. I'm confident about the vaccine. | - | X | Positive | 0.654 | 65% |
| I don't want to take COVID-19 vaccine. Flu kills more people than COVID-19. | - | X | Negative | 0.082 | 8% |
| I am not opposing but I will not take the vaccine yet. | X | - | Negative | 0.032 | 3% |
| I still want to be assured that COVID-19 vaccine is safe. | - | X | Negative | 0.225 | 22% |
| I cannot take COVID-19 vaccine because of my personal belief. | X | - | Negative | 0.239 | 24% |
| The pros outweigh the cons. So, I will take the vaccine. | X | - | Positive | 0.742 | 74% |
| I will take the COVID vaccine and overcome infections and deaths. | X | - | Positive | 0.671 | 67% |
| I will take the vaccine; I have chronic existing condition. | X | - | Positive | 0.484 | 48% |
| **Average Sentiments Score:** |  |  | **Combined Average:** | 0.457 | 46% |

The negative sentiments by some preschool workers (teaching and non-teaching staff) signaled distrust towards the COVID-19 vaccine, concerns about potential negative effects on pregnancy or women on lactation, and inadequate information about the vaccine. While the trust issue was borne out of concern that the vaccine was too new, the fears about some potential negative effects was a product of misinformation and conspiracy theories. On the other hand, the desire for more information offers insight into the need for adequate patient education and public briefings by the relevant public health agencies.

In addition to the shared negative opinions by all workers, the non-teaching employees further identified two (2) negative sentiments, two of which arose from genuine concerns. For example, one coded response indicated that some workers were not opposing the vaccine but needed more time to study the side effects. The second reason given by respondents for vaccine hesitancy in rejecting the vaccination was "personal belief." However, the nature and basis of the beliefs were unclear, whether based on religion or culture.

Similarly, the teaching staff expressed three major negative sentiments: (i). the need for further assurances that the COVID-19 vaccine is safe; (ii). Further observation of the vaccine outcomes in other people before committing to the vaccination; (iii). outright rejection of the vaccine based on misinformation or conspiracy theories. The results indicate the need for proper health education by the relevant public health agencies [4,50]. However, the third negative sentiment, which objected to the COVID-19 vaccine, argues that "flu kills more people than COVID-19," an expression that appears rooted in misinformation or conspiracy theory [51-53].

On the other hand, there were strong positive opinions about the COVID-19 vaccine that influence vaccine acceptance intentions posited by the workers. The rationale for the position sentiments includes the desire to "resume normal life," "helping to reach the herd immunity," protect themselves and family members against the coronavirus disease infection, and to be able to "visit family and friends again." Both categories of respondents also identified separate but related positive perceptions about the vaccine, as listed in Table 1. The only neutral opinion viewed the vaccine positively but remained nervous about accepting it due to safety fears. These sentiments correspond with conclusions from other studies on COVID-19 vaccine acceptance or hesitancy [52-53]. However, such fears can be addressed through adequate health education [4,50].

### 3.3 Association Between Employee Job Category and COVID-19 Vaccine Hesitancy

This Section examines the relationship between workers’ job category and COVID-19 vaccine hesitancy and helps to analyze the second research objective (RO2). The survey asked the employees about their intention to take the COVID-19 vaccine. In examining the responses, we categorized the employees into two groups: the teaching staff (teachers and classroom assistants) and the non-teaching/support staff. More support staff participated in the survey than teachers (66% to 34%; a ratio of nearly 2:1, respectively – Figure 2). The survey responses from the support staff show an evenly split between those accepting the COVID-19 vaccine (50%), while the hesitant group (outright rejection of vaccine and the undecided (36.4%, 13.6%, respectively). A slightly more teaching staff (53.8%) felt positive about the COVID-19 vaccine, compared to the vaccine hesitant teachers (30.8% outright rejection and 15.4% undecided), as indicated in Figure 2.

**Figure 2.**
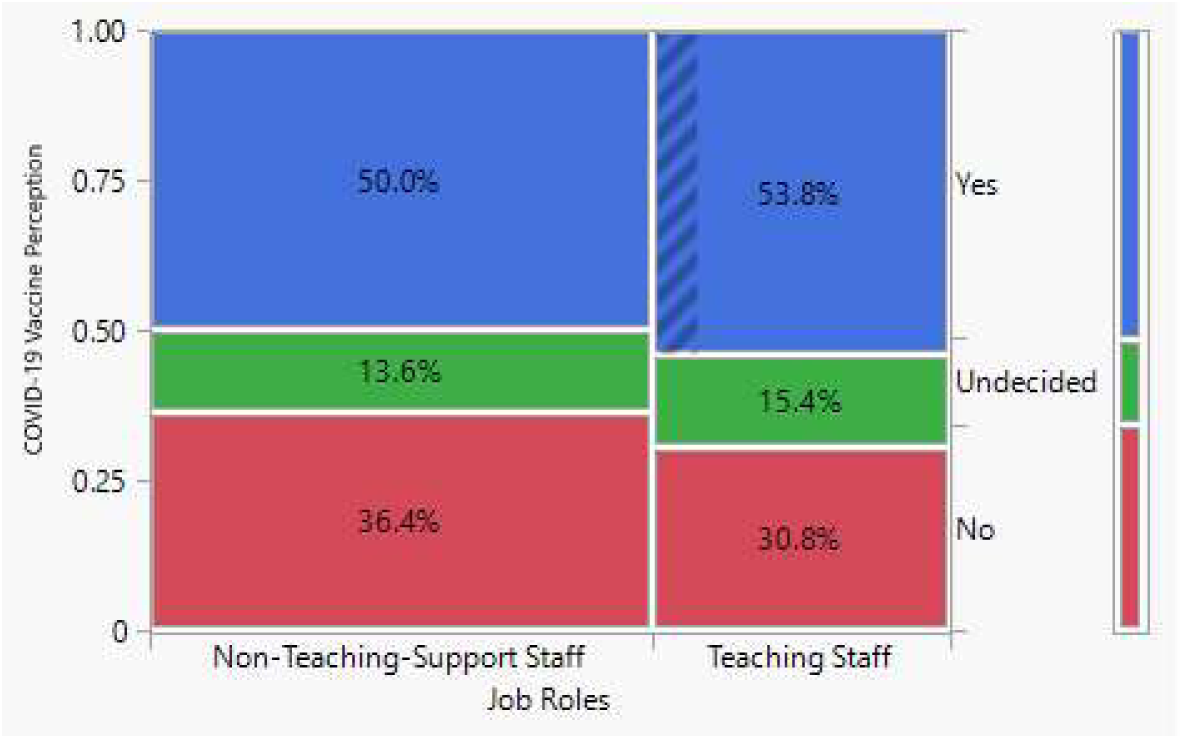
Employees’ perceptions and plans toward taking the COVID-19 vaccine. The ‘undecided’ and ‘no’ categories constitute the hesitancy group.

### 3.4 Does Employment Category Influence Vaccine Hesitancy?

Parts of the study examined the propositions about respondents’ type of employment (e.g., essential workers or frontline employees) on COVID-19 vaccination hesitancy. The employees who participated in the survey included school bus drivers, office staff, family advocates, classroom attendants, and teachers/support teachers. As explained earlier, employees such as teachers, support teachers, and bus drivers are employees who have the most direct contact with students and can be termed “frontline staff” [17]. In our analysis, we decomposed all the employees into two main categories, namely teaching and non-teaching/support staff (Figure 2).

The questionnaire asked the respondents about their intentions to take COVID-19 vaccine when offered. The respondents were required to select one of the three options, including "yes," "no," and "unsure." The options were re-classified into three options, one category being "hesitant," "no," or "unsure" answers. The positive responses were the "yes" answers, while the “no” and “unsure” options were interpreted as being hesitant. The study by Reno et al. [45]. on vaccine hesitancy in Italy adopted a similar re-classification of the survey responses.

The Chi-Square (χ^2^) test of independence was used to evaluate the association between employee categories (teaching v non-teaching/support staff) and vaccine hesitancy. The results show that slightly fewer teaching staff were hesitant at taking the COVID-19 vaccine than the support staff (46.2% < 50%). The Pearson Chi-square value was quite low at 0.009, with a significantly high p-value (p > 0.05). The results imply that the slight difference between the hesitant teachers (46.2%) compared to support staff (50%) was not significant, hence the employee category and vaccine hesitancy were independent. Thus, employment category among preschool workers does not influence COVID-19 vaccination intensions.

### 3.5 Factors Influencing Employees’ Vaccination Decisions

Table 2 presents different factors that influenced vaccination decisions among preschool employees. The stated factors come from the survey responses as explained above. In an open-ended question, the respondents were asked to give reasons for decisions made about their intention to accept or reject the COVID-19 vaccination when offered. Altogether, there were eighteen (18) identifiable reasons for the workers’ decisions on COVID-19 vaccination. Some respondents gave more than one factor. These factors are categorized as vaccine acceptance or hesitant. The hesitant category includes those that entertain fear or doubt about the vaccine, genuine or baseless [54].

**Table 2.** COVID-19 Vaccine Decision Reasons By COVID-19 Vaccine Accept-Hesitant.

| **Reasons for COVID-19 Acceptance versus Hesitancy** | **Accept**<br>**(Expected)** | **Hesitant**<br>**(Expected)** | **Total %** |
| --- | --- | --- | --- |
| Existing chronic health conditions | 2.86% (0.514) | 0.00 (0.486) | 2.86 |
| Confident about COVID-19 vaccine | 2.86% (0.514) | 0.00 (0.486) | 2.86 |
| Covid-19 Vaccine too new to trust | 0.00 (3.6) | 20.0% (3.4) | 20.00 |
| Eager to resume normal life | 5.71% (1.029) | 0.00 (0.971) | 5.71 |
| Flu kills more people than COVID-19 | 0.00 (0.514) | 2.86% (0.486) | 2.86 |
| Help reach herd immunity/stop pandemic | 8.57% (1.543) | 0.00 (1.457) | 8.57 |
| I am not opposed to getting it | 0.00 (0.514) | 2.86% (0.486) | 2.86 |
| I need more information | 2.86% (1.029) | 2.86% (0.971) | 5.71 |
| Nervous about side effects | 0.00 (1.029) | 5.71% (0.971) | 5.71 |
| Observing the outcomes in others | 0.00 (1.029) | 5.71% (0.971) | 5.71 |
| Personal belief | 0.00 (0.514) | 2.86% (0.486) | 2.86 |
| Pregnancy Related | 0.00 (1.029) | 5.71% (0.971) | 5.71 |
| Pros outweigh cons | 2.86% (0.514) | 0.00 (0.486) | 2.86 |
| Protect self, my family, and others | 8.57% (1.543) | 0.00 (1.457) | 8.57 |
| Stay safe and healthy | 5.71% (1.029) | 0.00 (0.971) | 5.71 |
| Still need assurance it's safe | 2.86% (0.514) | 0.00 (0.486) | 2.86 |
| Too many people have died | 2.86% (0.514) | 0.00 (0.486) | 2.86 |
| Visit family-friends again | 5.71% (1.029) | 0.00 (0.971) | 5.71 |
| Total %, (Expected Value) | 51.43% | 48.57% | 100% |
| $\chi^2=0.009$; $p>0.05$ | | | |

The most frequently reported decision factor was offered by respondents against the COVID-19 vaccination including “Covid-19 Vaccine too new to trust” (20%). The next most popular reasons offered by employees who favored the COVID-19 vaccine included “help reach herd immunity/stop pandemic” (8.57) and ‘protect themselves, family, and others (8.57%). Table 2 presents the full list. The listed factors were associated with COVID-19 vaccine hesitancy or acceptance respectively.

Further diagnostics of the factors for COVID-19 vaccine hesitancy compare the reasons offered by teachers versus non-teaching/support staff as revealed in Table 2. The analysis of the sentiments also reveals the following implications:

i. The employees who had positive sentiments also plan to take the COVID-19 vaccine and gave mainly scientific reasons to support their stance compared to those who opposed it. The explanations offered by the pro-vaccine group include "helping the community to achieve herd immunity and view vaccine as an effective way of staying safe." ^26^ Also, these respondents recognize that the "benefits of taking the vaccine outweigh the side effects." ^55^
ii. Some employees who did not want to take the vaccine also had genuine concerns, which, if addressed, will help them make a prudent decision. For example, some of the employees who disagreed with taking the vaccine voiced out about it not being long enough to trust its potency, while others wanted the observe the side effects on others before making a firm decision.
iii. Few respondents with opposing views about the vaccine gave non-scientific reasons, including "personal beliefs" or distrust of the vaccine for ‘not being around long enough. Other comments aligned more with COVID-19 vaccine misinformation. For example, reasons such as "trying to get or being pregnant" are not necessarily based on scientific evidence. Taking the COVID-19 vaccine does not negatively impact pregnancy [56-59].

## 4. Conclusion

This study provides evidence about the factors that influence COVID-19 vaccine hesitancy. The sentiment analysis using AML offers insights into why the respondents in our case involving the preschool workers were hesitant toward COVID-19 vaccination. The workers’ sentiments and perceptions about the COVID-19 vaccine in our case study corroborate the global experience. The attitude and behavior of workers towards COVID-19 vaccine identifies three categories of people: those in compliance, the opposers, and the unsure. The sentiment analysis results show that the preschool workers were generally neutral toward the vaccine. However, those who were neutral about the vaccine were also hesitant about taking the vaccination.

Further, there was no significant difference in workers’ vaccination intensions based on job categories (teachers versus support staff). However, few teaching staff (who hold college degrees) have strong negative sentiments rooted in misinformation or conspiracy theories, such as positing that "flu kills more people than COVID-19.

The above negative sentiments towards COVID-19 vaccine have several implications. First, the perceptions of preschool workers in this case study are not isolated but align with the global negative narrative from several other case studies from employees in different organizations. Second, some studies show that the more educated people are, the less hesitant they are toward the COVID-19 vaccine. Our study aligns with this conclusion in some parts. However, the results also show that educated people can be susceptible to misinformation and conspiracy theories, causing vaccine hesitancy and sometimes championing misinformation. Thirdly, employees who felt optimistic about the vaccine cited scientific reasons, while the hesitant group was primarily influenced by misinformation, fear, and distrust. Finally, the local health department needs to educate the public about the benefits of taking the COVID-19 vaccine, as even the educated can fall prey to misinformation and conspiracy theories, leading to vaccine hesitancy.

The results of the sentiment analysis also showed that some workers who were neutral or would not take the COVID-19 vaccine do so not necessarily because they oppose it but need more information or health education on the potency and validity of the vaccine. Interestingly, these factors identified in this study as positive, negative, or neutral sentiments toward COVID-19 vaccine among preschool workers corresponds with conclusions from many studies on COVID-19 vaccine hesitance or acceptance [51,52]. The study contributes to the literature on factors accounting for vaccine hesitancy among workers in educational institutions. Also, the methods used for the results analysis demonstrate the use of less-complex data analytics methods’ usefulness in evaluating textual data in healthcare and biomedical research.

### 4.1 Limitation of Study

The survey reported in this study was conducted in Feb/Mar 2021, when the COVID-19 vaccination schedule began for the most vulnerable elderly population. There is a potential lag between the survey responses obtained then and the current reality in 2024. Also, most of the respondents in this survey belonged to the younger population and were not yet assigned to take the COVID-19 vaccines. We intend to conduct a follow-up survey to learn if those with negative sentiments later changed their position to accept the vaccine and vice versa. Based on the preschool organization policy and to ensure anonymity of the survey, workers’ personal information capable of identifying the respondents was not collected. This explains why there are no discussions about specific age group(s) or gender, or respondent’s field of study.

Furthermore, the sample size of 30 obtained in this study may not appear large. However, bearing in mind that this was a single early learning center operating as a small business, nearly all the employees participated willingly in the survey.

Future study will also survey preschool students and their parents to understand the current perceptions about the COVID-19 vaccine.

## Data Availability

The authors do not have the authority to share the data.

## Human Subjects Approval Statement

The study was part of a consulting project requested by the preschool organization. Necessary approvals were granted by the preschool organization.

## Conflict of interest

The authors declare no conflicts of interest

## Notes

### Competing Interest Statement

The authors have declared no competing interest.

### Funding Statement

This study did not receive any funding.

